# Drug-Related Problems and associated factors among hospitalized pediatric patients at the University of Gondar Comprehensive and Specialized Hospital

**DOI:** 10.1101/2022.09.12.22279865

**Authors:** Atalay Amsalu, Adhanom Gebreegziabher Baraki, Esileman Abdela Muche

## Abstract

**Introduction:** Drug-related problem is any event involving drug therapy that interferes with a patient’s desired clinical outcome. Hospitalized pediatric patients are particularly prone to drug-related problems. Thus, this study aimed to assess drug-related problems and associated factors among patients admitted to the pediatric ward of the University of Gondar Comprehensive and Specialized Hospital, Ethiopia.

**Methods:** A hospital-based cross-sectional study was conducted among pediatric patients admitted to the University of Gondar Comprehensive and Specialized Hospital from May 1, to July 30, 2021. A Simple random sampling technique was employed to select study participants. Odds ratio with 95% confidence interval was computed for each variable for the corresponding P-value to see the strength of association. Those variables which have a P-value of < 0.25 in Bivariable analysis were entered in Multivariable analysis to determine factors associated with drug related problems.

**Results:** A total of 180 drug-related problems were identified in 145 participants with an overall prevalence of 40.2% [95% CI (35.5-45.4)]. Dose too low (35.56%), needs additional drug (28.89%) and dose too high (21%) were the commonest type of drug-related problems identified. The most important interventions made were dose adjustment (52%) and the addition of drugs (30%). The presence of comorbidity [AOR = 3.32, 95% CI (1.88-5.88)], polypharmacy [AOR = 4.22, 95% CI (2.21-8.10)], and more than 6 days stay in the hospital [AOR =7.59, 95% CI (3.76-15.33)] were independent predictors for the occurrence of drug-related problems.

**Conclusion:** Drug-related problems are common among hospitalized pediatrics at the University of Gondar Comprehensive and Specialized Hospital. The presence of comorbidity, polypharmacy and prolonged hospital stay were predictors of drug-related problems. Therefore, health care providers have to work in collaboration with clinical pharmacists and give due attention to those patients with comorbidity, polypharmacy and patients who stayed longer days in the hospital.

## Introduction

Drugs are very vital tools in medical practice contributing to the improvement in the quality and expectancy of patients’ life by alleviating symptoms, halting disease progression, preventing transmission, and curing diseases but they may lead to emergency hospital visits, increase hospitalizations, in-patient, and outpatient care complications when used inappropriately (1-3).

A drug-related problem (DRP) is defined as an undesirable event involving drug therapy that actually or potentially interferes with desired health outcomes and requires professional judgment to resolve it. DRPs are classified into seven categories as unnecessary drug therapy, needs additional drugs, ineffective drugs, dose too low, dose too high, adverse drug reactions, and noncompliance(4). Drug related problems can originate at any process of medication use (prescribing, transcribing and verifying, dispensing, administering, and monitoring and reporting(5).

The pediatric medication use process is complex and error-prone because of the multiple steps required in calculating, verifying, preparing, and administering doses that complicate the course of their diseases and cause treatment failure and adverse drug reactions(6). Hospitalized pediatric patients are also often exposed to an extensive number of medications especially those with longer stays which lead to drug-related problems (7). Children are estimated to be three times more than adults to experience harm related to their medication (8).

DRP causes significant mortality, morbidity, and also an economic crisis in the health care system. Drug related problems as a cause to hospital admission ranging from 2% to 10.3% while in admitted patients ranges from 27.8% to as high as 81% and about 22% of patients are discharged with DRP (9, 10). A review of observational studies in Europe concluded that approximately 3.6 % of hospital admissions are caused by adverse drug reactions, and up to 10 % of patients experience an adverse drug reaction during their stay in the hospital(11).

The estimated annual cost of drug-related morbidity and mortality resulting from nonoptimized medication therapy was $528.4 billion in 2016(12) in United states of America, in Japan US$799,966.6(13), in Australia £100,707(14) and in Nigeria, 1.83 million naira (USD 15,466.60) was spent to treat all the patients admitted due to DRPs from 2006 to 2007(15).

A cohort Study conducted on DRPs among hospitalized Children in Saudi Arabia showed that the incidence of DRP was 35.9%(16). Another cross-sectional study done in Zewditu memorial hospital Ethiopia showed the prevalence of drug-related problems was 31.57(17). A cohort study conducted in Jimma medical center concluded that the incidence of DRP was 48.8%(18). The major classes of drugs involved in the DRPs were anti-infectives (19, 20). Studies showed that polypharmacy, type of medical conditions, type of admission, length of hospital stay, and number of medical conditions were the factors that were associated with DRP among pediatric patients(21-23).

The assessment and identification of DRPs, care plans, and follow-up care are the major ways to reduce the problems(24). Systematic reviews and metanalysis done concluded that pharmacist interventions are effective for reducing DRPs in hospitalized pediatric patient (25, 26). Another cross-sectional study conducted in Malaysia showed 81.8% of interventions were accepted (27).

Another cross-sectional study conducted in Cote d’Ivoire hospitalized pediatrics showed the acceptance rate of pharmaceutical interventions was 94.8%(19). A cohort study was done in Addis Ababa showed 92.15% were fully accepted, 3.72% partially accepted, and 4.13% were not accepted (28).

Pediatrics are special populations that need special attention in their drug therapy but they have been suffered from this problem. The magnitude of DRPs and its predictors was not known among pediatric admitted patients in the study setting. Therefore, this study was aimed to assess the prevalence of DRPs and factors associated with DRPs at the pediatric wards of University of Gondar Comprehensive and Specialized Hospital, Ethiopia.

## Methods

### Study design

A hospital-based cross-sectional study was conducted at the pediatrics wards of the University of Gondar Comprehensive and Specialized Hospital.

### Study area and period

The study was conducted at the pediatrics wards of the University of Gondar Comprehensive and Specialized Hospital for a period of 3 months from May 1, to July 30, 2021.

### Population

All admitted pediatric patients less than 15 years of age were the source population. Those Pediatric patients who were admitted to the pediatric inpatient wards during the study period, and who fulfilled the inclusion criteria were the study populations

### Eligibility criteria

#### Inclusion criteria

- Patients less than 15 years of age and admitted to the pediatric ward for 24hrs during the study period.
- Those parents who gave informed consent and assent of pediatrics to participate in the study were included

#### Exclusion criteria

- Critically ill patients requiring intensive care unit (ICU) admission were excluded.
- Oncologic pediatric patients were also excluded

#### Sample size determination and sampling technique

The sample size was determined by using single proportion formula assuming the prevalence of DRP to be 32%(17) (p=0.32) to get the minimum sample size and with a 95% confidence interval (α=5%) as follows.

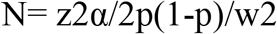

Where:

N=sample size, z critical value=1.96, p=q=0.32 W= precision (marginal error) =0.05

Then substitution of values for each:

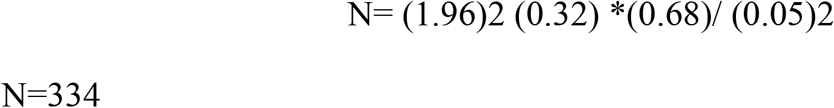

By adding 10% of the calculated sample size to compensate for non-respondents the final sample size was 368.

So, the final sample size was selected with the highest value which was obtained from the outcome variable and 368 study participants were included in the study.

The study participants were selected by simple random sampling method using bed numbers as sampling frame using lottery method until the required sample size for the study was obtained.

#### Study variables

In this study, the dependent variable was a DRP. sociodemographic character of the patient (sex, age, weight, body mass index, MUAC), medication fee, disease type, nutritional status, length of treatment, comorbidity, number of drugs and disease, drug dosage regimen, history of immunization, history of allergy, history of prior admission and laboratory values were the independent variables.

#### Operational definitions

- Drug-related problem – according to Cipolle, DRP is any undesirable event experienced by a patient which involves, or is suspected to involve drug therapy, and that interferes with achieving the desired goals of therapy and requires professional judgment to resolve. It includes unnecessary drug therapy, need for additional drug therapy, adverse drug reaction, ineffective drug inappropriate dosage, adverse drug reaction and noncompliance(4).
- Unnecessary drug therapy-is A DRP that occurs when there is no valid medical indication for the drug at the time, or multiple drug products are used when only single-drug therapy is appropriate, or the condition is best treated with nondrug therapy(4).
- Need for additional drug therapy-is a DRP that occurs when there is a medical condition needing new drug therapy, or preventive therapy is needed to reduce the risk of developing a new condition, or a medical condition requires combination therapy for better efficacy(4).
- Ineffective drug-is a DRP where the drug is not the most effective for the medical problem, or the condition is refractory to the drug product being used, or the dosage form is inappropriate(4).
- Inappropriate dosage-refers to dosages both too low and too high(4).
  - Dosage too high—is a DRP where the dose is too high or the dosing frequency is too short or the duration of therapy is too long for the patient, or a drug interaction causes a toxic reaction to the drug product, or the dose was administered too rapidly.
  - Dosage too low-is a DRP that occurs when the dose is too low to produce the desired outcome, or the dosage interval is too long, or a drug interaction reduces the amount of active drug available, or the duration of therapy is too short.
- Noncompliance-is a DRP that occurs when the patient does not understand the instructions, or the patient prefers not to take or forgets to take the medication, or the cost of the drug product is not affordable for the patient, or the patient cannot swallow or self-administer the medication properly, or the drug product is not available for the patient(4).
- Adverse drug reaction _ “an appreciably harmful or unpleasant reaction, resulting from an intervention related to the use of a medicinal product, which predicts hazard from future administration and warrants prevention or specific treatment, or alteration of the <u>dosage regime</u>n, or withdrawal of the product.” (29)
- Comorbidity_ is considered when any distinct additional entity that has existed or may occur during the clinical course of a patient who has the index disease under study (30).
- Potential drug-drug interactions-are those that could theoretically take place when two or more drugs are prescribed to a patient (31).
- Polypharmacy - taking more than 4 medications during their hospitalization (17, 32)
- Medication adherence - “the extent to which a patient acts following the prescribed interval, and dose of a dosing regimen(33). Adherence was assessed using a medication adherence rating scale. it is a 10-item self-report instrument in which a yes/no response is given to questions asked. The total scores range from 0 (low likelihood of medication adherence) to 10 (high likelihood). The patient is adherent is when the score is greater than 5 (34)
- Pediatrics: are those age groups less than 15 years including neonates (from birth to 28 days), infants (1 month to 1 year), toddler (1 year to 3 years), preschool (3 years to 5 years), school-age (5 years to 10 years)and adolescent (10 to 15 years) (35).

#### Data collection and management

##### Data collection tools

Relevant information regarding patient’s demographics and clinical characteristics (e.g., diagnosed diseases, comorbidities, history of allergies, vital signs, pertinent laboratory and diagnostic tests, relevant past medical and medication history, current medication including OTC medications, length of stay) were collected by two clinical pharmacists by using data abstraction format from a patient chart and by interviewing patients.

##### Assessment of Adherence

Medication non-adherence is a major issue in hospitalized patients. Adherence was assessed by using a medication adherence rating scale (MARS). It is A 10-item self-reported Adherence measurement that consists of ten questions with closed dichotomous (yes / no) answers. Adherence is achieved when the result is greater than five according to this tool (36).

##### Data collection procedure

Two clinical pharmacists were trained as data collectors by the principal investigator extensively for one day about the objective of the study, methods of data collection including data extraction from patient charts as well as techniques of interviewing patients, data handling, ethical approaches, and DRP identification.

First; caregivers were asked for voluntary interviews and participation in the study. The medical record number of each patient was used to avoid duplication and keep confidentiality.

##### Identification of DRPS

Drug related problems were classified according to Cipolle, Morley, and Strand’s DRPs method with a slight modification. DRPs were identified by using Nelson textbook of pediatric 21th edition(37), pocket book of pediatric hospital care 2013(38), SAM guideline of Ethiopia 2019, and other updated guidelines by evaluating the appropriateness of medication in terms of indication, dosage, effectiveness, and safety. Patients’ clinical characteristics were taken into account when deciding about the appropriateness of the dosage regimen. The recommendations were done by a team of experts and forwarded to physicians. Drug-drug interaction was checked by using Medscape application drug interaction cheeker drug reference V1096.0.

Adverse drug reactions were identified from patient or caregiver interviews and investigating patient data for any possible adverse reactions related to patients’ medications.

To assess the causality and severity of the suspected adverse drug reactions, the Naranjo scale of causality and Hartwig severity assessment scale were used respectively (39, 40).

##### Data quality control technique

The data collection tool was pretested in 19 patients (5% of the sample size) admitted to the pediatric ward of Felege Hiwot Comprehensive and Specialized Hospital to check for uniformity and understandability of the checklist after which modification for its appropriateness and suitability was done. The data of the pretest was not included in the study. The supervisor (one clinical pharmacist other than data collectors) supervised the data collection process and give feedback and correction on daily basis by checking the quality, accuracy, consistency, and completeness of the data and asserted by signatures of data collectors and the supervisor. After data collection, the data was coded, edited, and cleaned to ensure accuracy, consistency, and completeness and entered into Epi data manager version 4.6 then exported and analyzed using SPSS version 21 soft were.

The quantitative and qualitative data were compiled and interpretation was done according to the finding of the study.

##### Data processing and analysis

The collected data was cleaned and checked for completeness and consistency before processing. Data were then entered into epi data version 4.6 and analyzed using Statistical Package for Social Studies (SPSS) version 21. The distribution of data was checked by using the Hosmer lemeshow goodness of fit test. Odds ratio (OR) with 95% confidence interval was also be computed for each variable for the corresponding P-value to see the strength of association. Those variables which have a P-value of < 0.25 in the bivariable logistic regression analysis were entered in Multivariable analysis to determine factors significantly associated with DRPs. Variables which have a P-values of <0.05 in multivariable regression analysis were considered as significantly association with DRPs.

The result of the analysis was presented by using frequencies, mean and standard deviation, texts, tables, and graphs. Finally, based on the results interpretation, comparison, conclusion, and recommendations were drawn.

#### Ethical consideration

Ethical clearance was obtained from the ethical review committee of department of clinical Pharmacy, Gondar University, and permission was also obtained from University of Gondar Comprehensive and Specialized Hospital and unit officials to conduct this study. Written consent was obtained from caregiver and verbal assent was given to study participants. Study participants or caregivers were informed about the purpose of the study and their participation was voluntary. The Participant assured that lack of willingness to involve in the study wouldn’t affect the service they get. Privacy of participants was ensured since patients are differentiated with their card number (no name) only. All information obtained from the participants was kept confidential and the data obtained was used for the research purpose only.

## Result

### Socio-demographic characteristics of participants

In this study, a total of 368 were selected to conduct study. Seven patients were excluded because of unwillingness to participate in the study and the response rate was 98.1%. More than half of the participants were male 199 (55.1%). The mean (± SD) age of study participants was 3.73 ± 1.48 years, the majority (24.7%) lie within the age group of 1-3 years. Moreover, the mean (± SD) weight and height of patients were 15.6 ±9.9 kg and 98.5 ±31.4cm) respectively (Table 1)

**Table 1:** Socio-demographic characteristics of pediatric patients attending at pediatric ward of UoGCSH, Ethiopia, May 1 to July 30, 2021

| Variables | Category | Frequency (N=361), (%) | mean ± SD |
| --- | --- | --- | --- |
| Age (years) | neonate (< 28 days) | 9 (2.5) | $3.73 \pm 1.48$ yrs. |
|  | Infant (29 days -1 year) | 85 (23.5) |  |
|  | Toddler (1-3 years) | 89 (24.7) |  |
|  | Preschool (3 -5 years) | 47 (13) |  |
|  | School age (5– 10 years) | 72 (19.9) |  |
|  | Adolescents (10 – 15 years) | 59 (16.3) |  |
| Gender | Male | 199 (55.1) |  |
|  | Female | 162 (44.9) |  |
| Residence | Rural | 231 (64) |  |
|  | Urban | 130 (36) |  |
| Source of medication | Health insurance | 284 (78.7) |  |
|  | paid | 77 (21.3) |  |
| Weight (kg) | <5 | 29(8) |  |
|  | 5-9.9 | 90(24.9) | 15.6 ± 9.9 kg |
|  | 10-14.9 | 81(22.4) |  |
|  | 15-19.9 | 43(11.9) |  |
|  | 20-24.9 | 44(12.2) |  |
|  | >25 | 74(20.5) |  |

UoGCSH=University of Gondar Comprehensive and Specialized Hospital

### Clinical data of participants

The majority of study participants had two disease conditions accounted for 135(37.4%). Most of the patients had 3 days of hospital stay accounted for 162 (44.9%) with the mean (± SD) duration of hospital stay was 4.65± 2.72 days. Regarding immunization status, 253 (70.1%) patients were fully immunized. The majority of the patients 291 (80.9%) had received less than 5 drugs per day. Patients’ level of adherence was assessed using interview and was 100% adherent. In addition, reconciliation was made between the medication order sheet and medication administration sheet to confirm adherence level and to assess whether there is a discrepancy. Overall, no difference between the medication order sheet and the administration sheet strengthens our result.

Among the prescribed drug classes, anti-infectives 576(51.3%) were the commonly prescribed drug class. A total of 116 drug-drug interactions were identified in 66 (18.3%). Among the identified drug-drug interactions, 71(61.2 %) of drug-drug interactions were pharmacokinetic interactions. In terms of severity of drug interaction 70(60.35%) were minor interactions. None of the patients has a prior history of known allergy (Table 2).

**Table 2:**
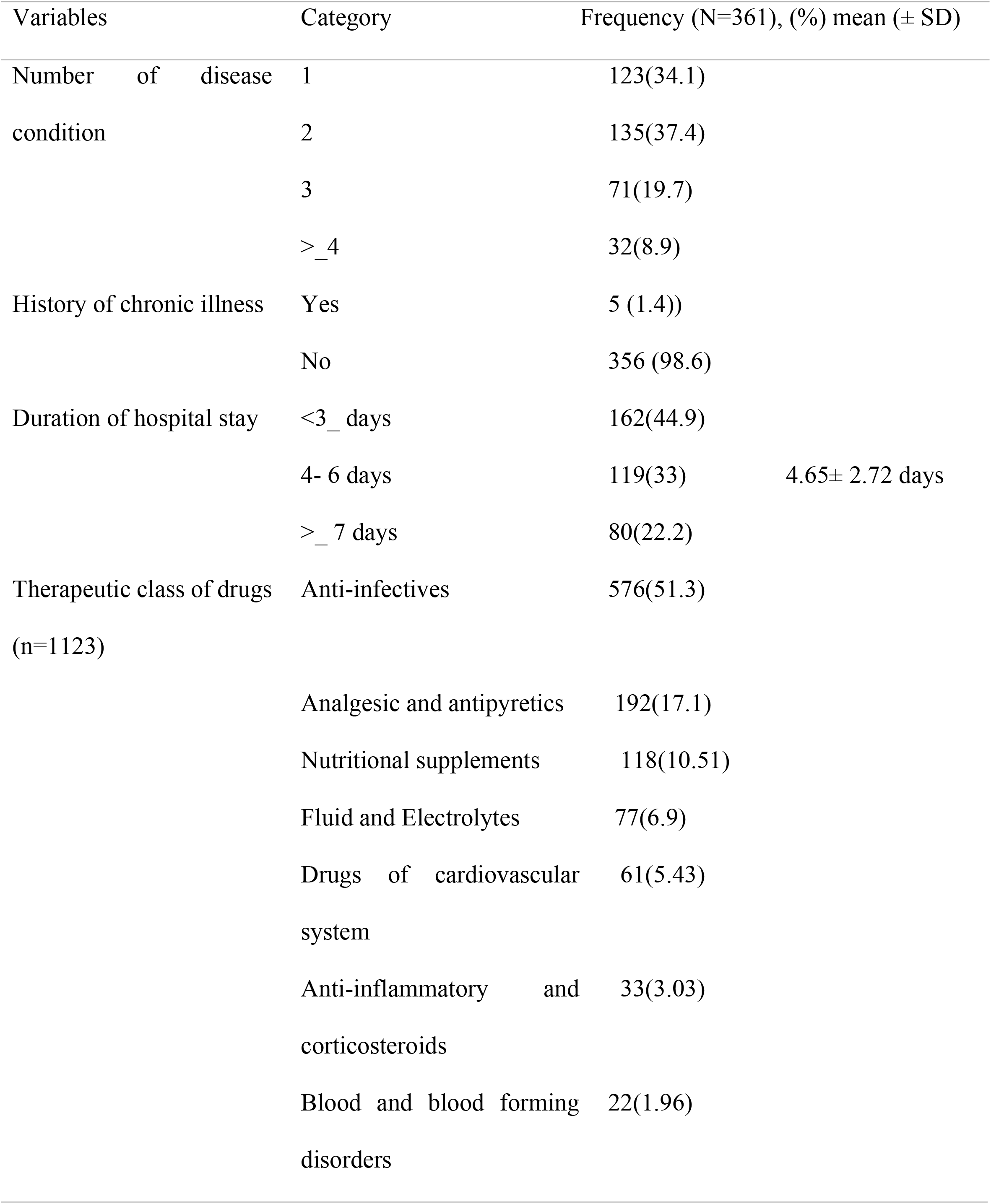
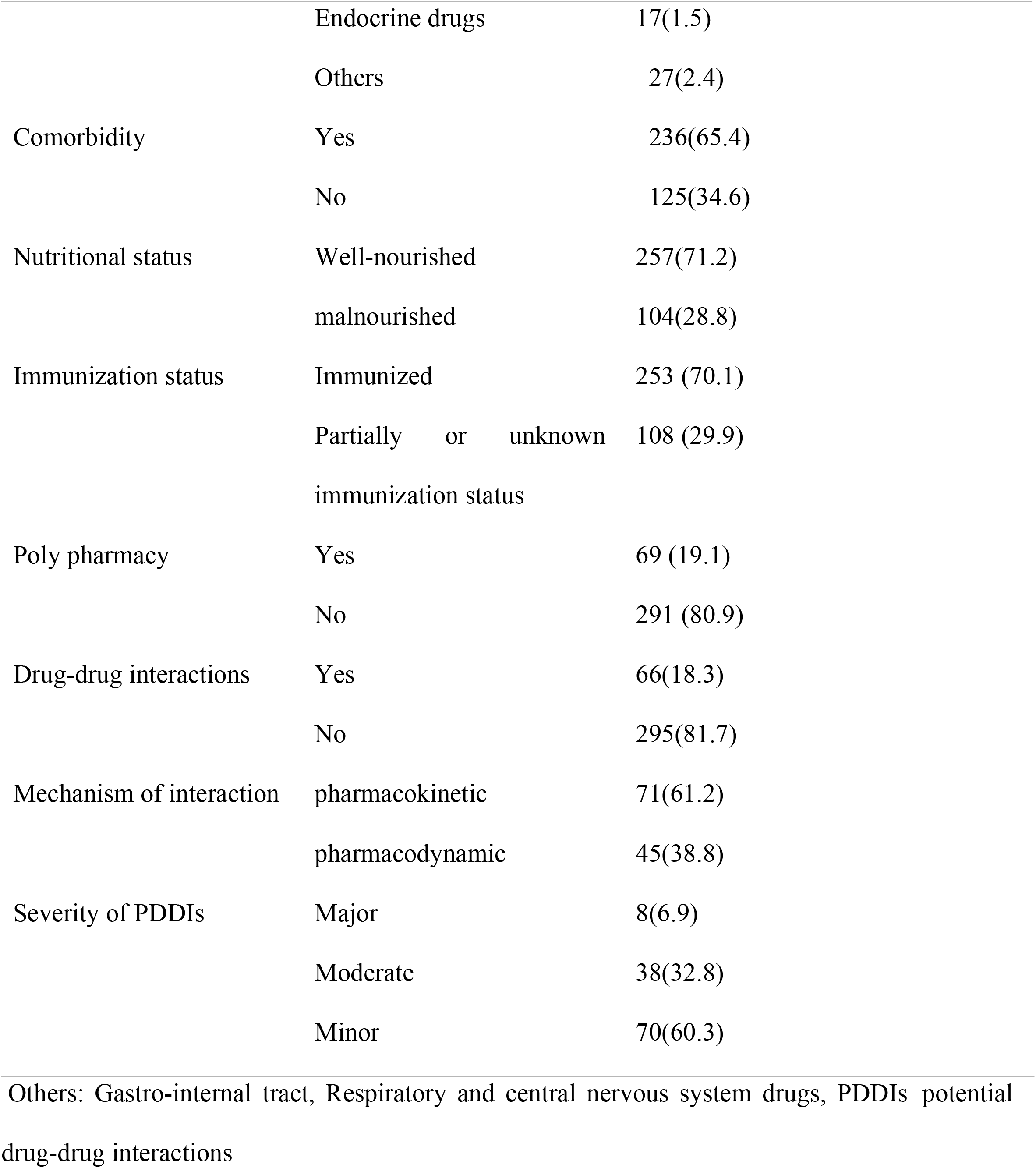
Clinical characteristics of pediatric patients attending at pediatric ward of UoGCSH, Ethiopia, May 1 to July 30, 2021

| Variables | | | Category | Frequency (N=361), (%) mean (± SD) | |
| --- | --- | --- | --- | --- | --- |
| Number of disease condition |  |  | 1 | 123(34.1) |  |
|  |  |  | 2 | 135(37.4) |  |
|  |  |  | 3 | 71(19.7) |  |
| | | | $\geq 4$ | 32(8.9) | |
| History of chronic illness |  |  | Yes | 5 (1.4)) |  |
|  |  |  | No | 356 (98.6) |  |
| Duration of hospital stay | | | $<3$ days | 162(44.9) | |
| | | | 4- 6 days | 119(33) | 4.65± 2.72 days |
| | | | $\geq 7$ days | 80(22.2) | |
| Therapeutic class of drugs (n=1123) |  |  | Anti-infectives | 576(51.3) |  |
|  |  |  | Analgesic and antipyretics | 192(17.1) |  |
|  |  |  | Nutritional supplements | 118(10.51) |  |
|  |  |  | Fluid and Electrolytes | 77(6.9) |  |
|  |  |  | Drugs of cardiovascular system | 61(5.43) |  |
|  |  |  | Anti-inflammatory and corticosteroids | 33(3.03) |  |
|  |  |  | Blood and blood forming disorders | 22(1.96) |  |

|  |  |  |
| --- | --- | --- |
|  | Endocrine drugs | 17(1.5) |
|  | Others | 27(2.4) |
| Comorbidity | Yes | 236(65.4) |
|  | No | 125(34.6) |
| Nutritional status | Well-nourished | 257(71.2) |
|  | malnourished | 104(28.8) |
| Immunization status | Immunized | 253 (70.1) |
|  | Partially or unknown immunization status | 108 (29.9) |
| Poly pharmacy | Yes | 69 (19.1) |
|  | No | 291 (80.9) |
| Drug-drug interactions | Yes | 66(18.3) |
|  | No | 295(81.7) |
| Mechanism of interaction | pharmacokinetic | 71(61.2) |
|  | pharmacodynamic | 45(38.8) |
| Severity of PDDIs | Major | 8(6.9) |
|  | Moderate | 38(32.8) |
|  | Minor | 70(60.3) |

Others: Gastro-internal tract, Respiratory and central nervous system drugs, PDDIs=potential drug-drug interactions

### Prevalence and nature of DRPs

Among the study participants, a total of 180 DRPs were identified in 145 patients and an overall prevalence was 40.2% (95% CI 35.5-45.4), a mean of 1.24 ± 0.50 per patient.

Among the identified DRP types, Dose too low was the most frequent DRP accounting for 64 (35.56%), needs additional drug was the second accounting for 52 (28.89%), and dose too high was the third 37 (21%) (Figure 1). Among the study participants who encountered DRPs, 114 (31%) of them have one drug related problem, 28(8%) have two and 3(1%) have three drug related problems respectively.

**Figure 1:**
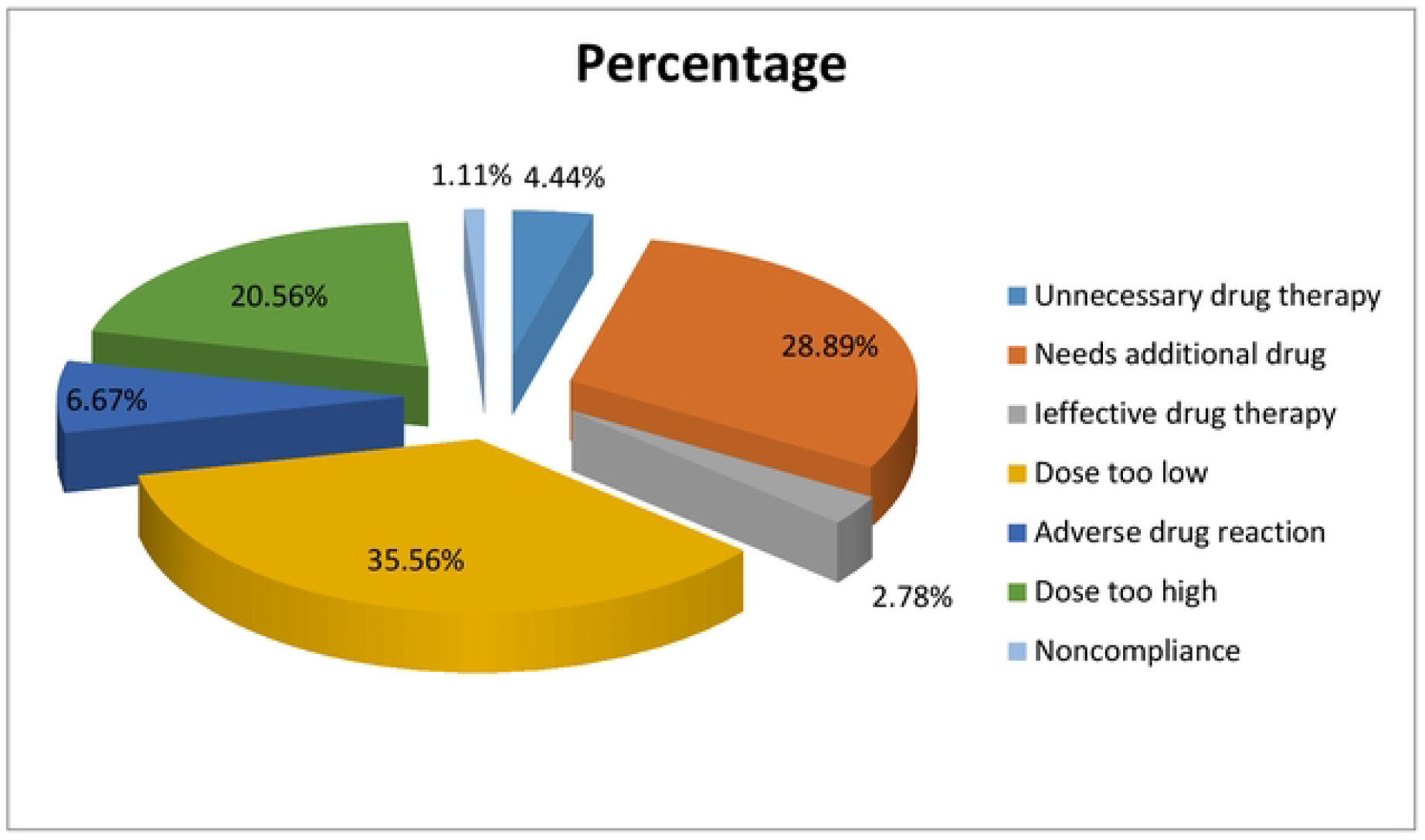
Type of drug related problems identified among study participants in the pediatric ward of UoGCSH, Ethiopia, May I to July 30, 2021.

### Diseases involved in DRPs

Using the WHO-ICD 10 classifications for a disease diagnosed, the most prevalent specific diseases implicated in DRPs were pneumonia 35 (19.4%), SAM 30 (16.7%) (Table 3)

**Table 3:** top ten diseases involved in DRPs at pediatric ward of UoGCSH, Ethiopia, May 1 to July 30, 2021

| S.N | Diseases | Frequency (%) |
| --- | --- | --- |
| 1 | Pneumonia | 35(19.5) |
| 2 | Severe acute malnutrition | 30(16.7) |
| 3 | Sepsis | 17(9.4) |
| 4 | Meningitis | 16(9) |
| 5 | Malaria | 6(3.3) |
| 6 | Post streptococcal glomerulonephritis | 6(3.3) |
| 7 | Tuberculosis | 6(3.3) |
| 8 | Heart failure | 6(3.3) |
| 9 | Intestinal parasite | 6(3.3) |
| 10 | Acute tonsillopharyngitis | 5(2.8) |

### Drugs and drug classes involved in DRPs

Using the WHO-ATC classification system for medications, the most frequently involved drug class in the DRPs were anti-infectives 111/180(61.7%) and analgesic–antipyretics 24(13.33%), vitamins and other nutritional supplements 22(12.22%) (Table 4).

**Table 4:**
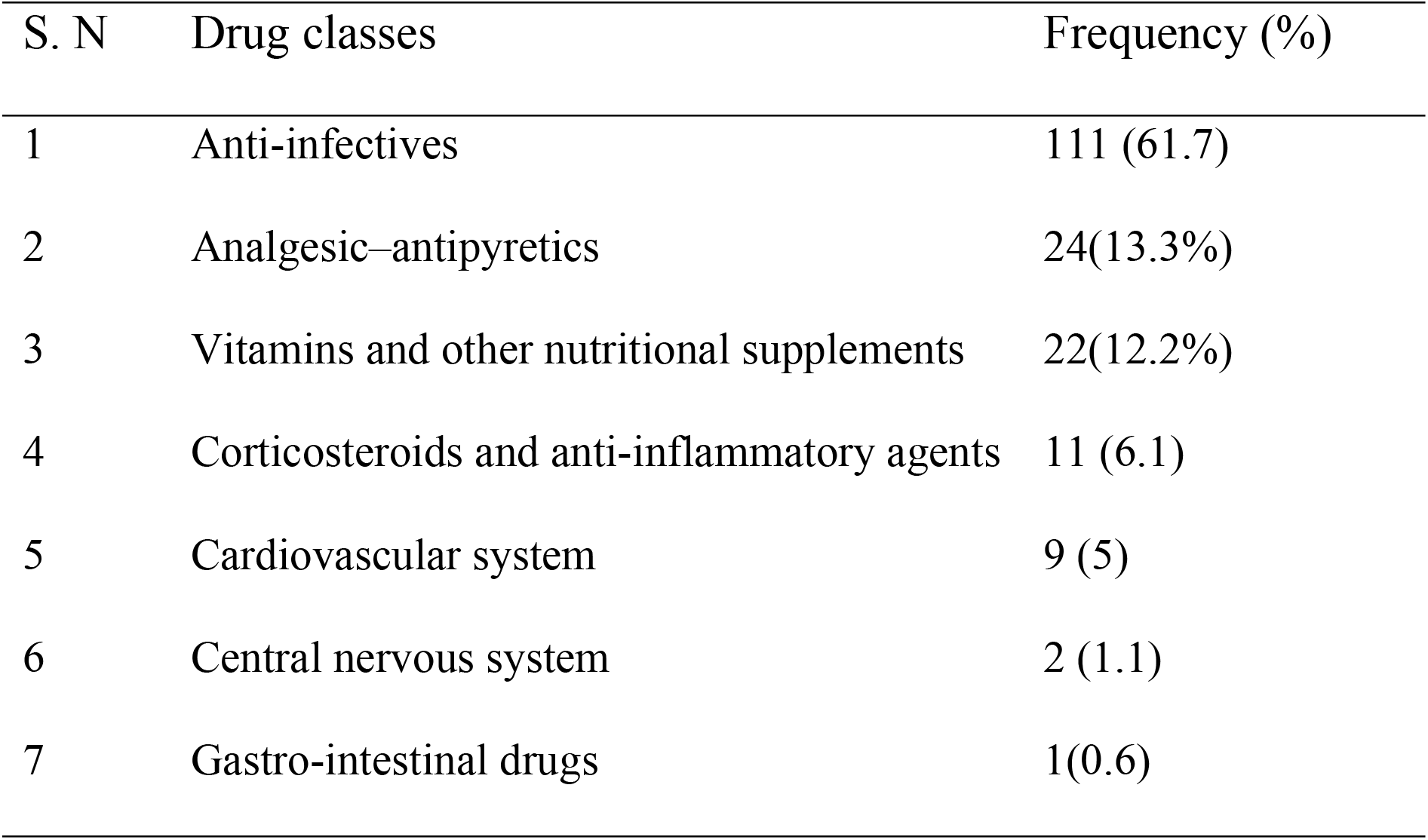
Drug classes involved in DRPs at pediatric ward of UoGCSH, Ethiopia, May 1 to July 30, 2021

| S. N | Drug classes | Frequency (%) |
| --- | --- | --- |
| 1 | Anti-infectives | 111 (61.7) |
| 2 | Analgesic–antipyretics | 24(13.3%) |
| 3 | Vitamins and other nutritional supplements | 22(12.2%) |
| 4 | Corticosteroids and anti-inflammatory agents | 11 (6.1) |
| 5 | Cardiovascular system | 9 (5) |
| 6 | Central nervous system | 2 (1.1) |
| 7 | Gastro-intestinal drugs | 1(0.6) |

From the prescribed drugs, the most frequently implicated drug was ceftriaxone 30(16.7%), paracetamol 15(10.34%), and Ready to use therapeutic feeding 14(9.7%) (Table 5).

**Table 5:**
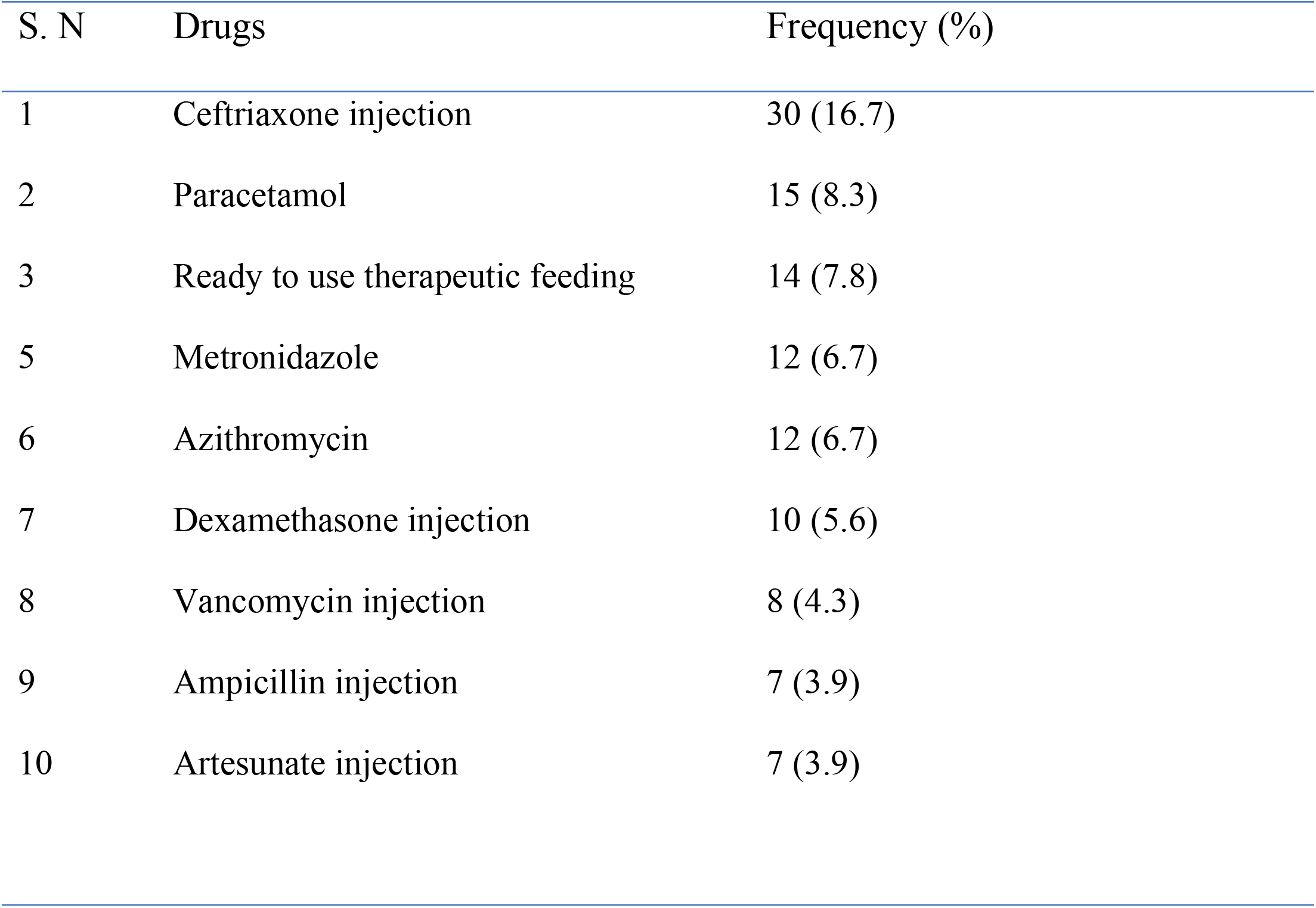
Top ten drugs involved in DRPs at pediatric ward of UoGCSH, Ethiopia, May 1 to July 30, 2021

### DRP type and its causes

Among the identified DRPs, Dose too low was the most frequent DRP accounting for 64 (35.56%), needs additional drug was accounting for 52 (28.89%), and dose too high the 37 (21%). Among the possible causes of DRPs, 51.7% were related to the wrong drug dose, a medical condition requires the initiation of drug 14%, and to attain synergistic effect needs additional drug therapy (10%) and drug interaction (10%) most frequent causes (Table 6).

**Table 6:**
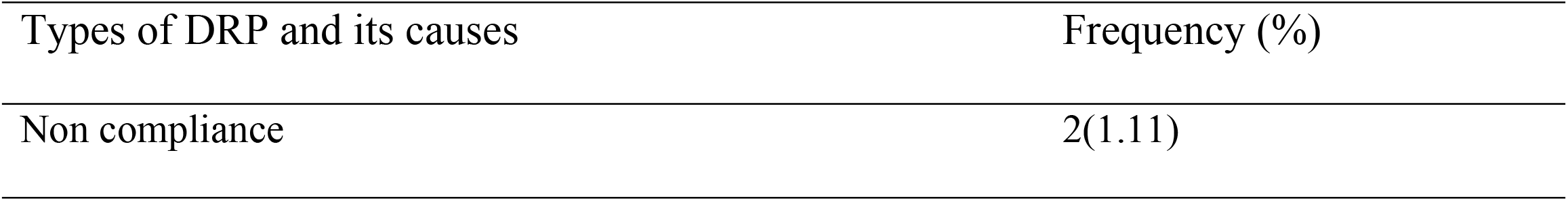
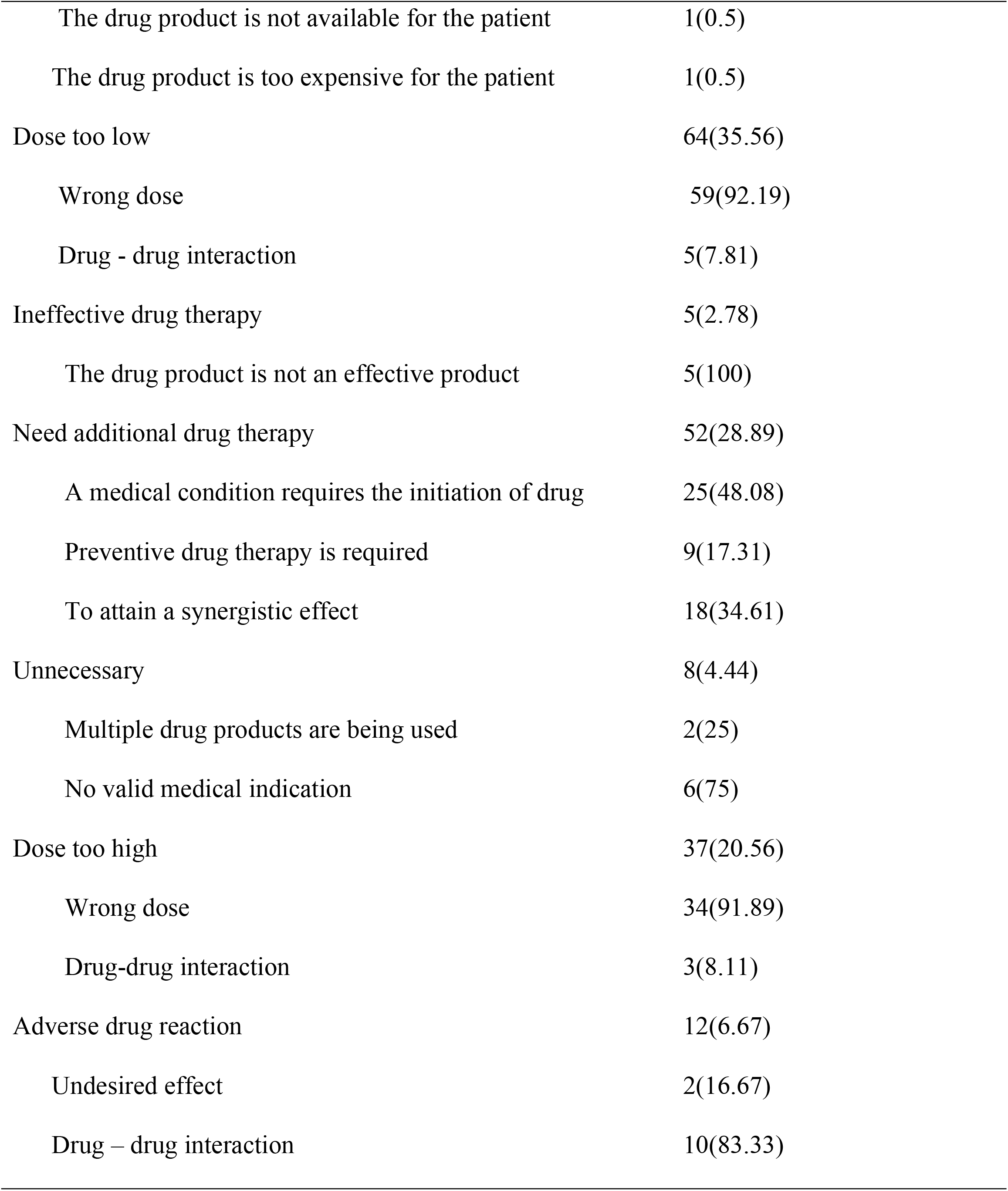
The common causes of DRPs identified among study participants at pediatric ward of UoGCSH, Ethiopia, May 1 to July 30, 2021 (N=180)

| Types of DRP and its causes | Frequency (%) |
| --- | --- |
| Non compliance | 2(1.11) |
| The drug product is not available for the patient | 1(0.5) |
| The drug product is too expensive for the patient | 1(0.5) |
| Dose too low | 64(35.56) |
| Wrong dose | 59(92.19) |
| Drug - drug interaction | 5(7.81) |
| Ineffective drug therapy | 5(2.78) |
| The drug product is not an effective product | 5(100) |
| Need additional drug therapy | 52(28.89) |
| A medical condition requires the initiation of drug | 25(48.08) |
| Preventive drug therapy is required | 9(17.31) |
| To attain a synergistic effect | 18(34.61) |
| Unnecessary | 8(4.44) |
| Multiple drug products are being used | 2(25) |
| No valid medical indication | 6(75) |
| Dose too high | 37(20.56) |
| Wrong dose | 34(91.89) |
| Drug-drug interaction | 3(8.11) |
| Adverse drug reaction | 12(6.67) |
| Undesired effect | 2(16.67) |
| Drug – drug interaction | 10(83.33) |

### The type of interventions provided and acceptance of interventions on DRPs

The intervention was provided for 98.9% of all the identified problems except one because of the problem was identified and intervened by a physician. 76.54% of the intervention were accepted and fully applied and 23.9% of interventions were not accepted and not applied. The major interventions provided were dose modification (52%) and addition of drugs (30%) (Figure 2).

**Figure 2:**
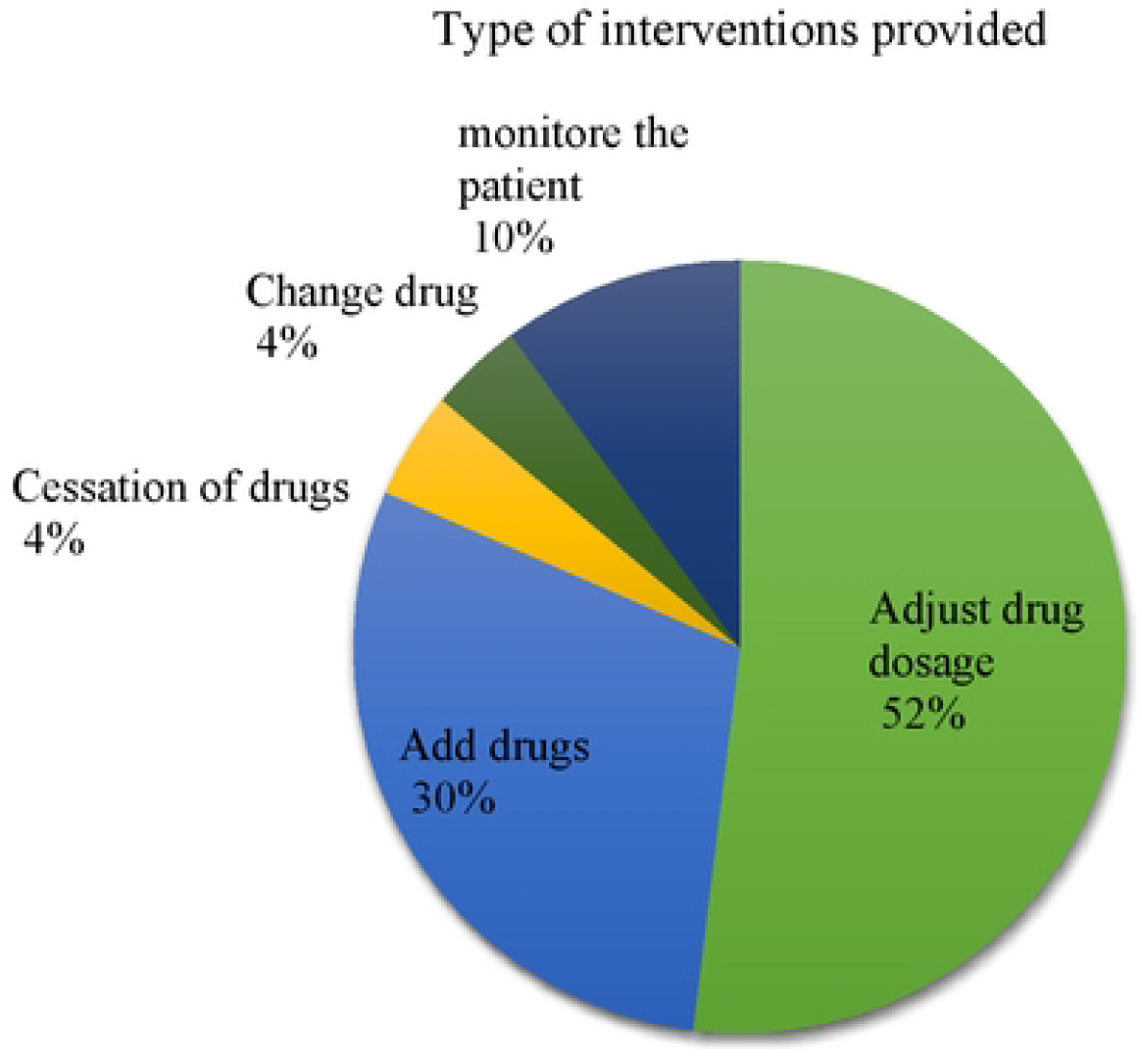
The type of interventions provided for the identified drug related problen1s at the pediatric ward ofUoGCSH, Ethiopia, May I to July 30, 2021.

### Factors associated with DRPs

Among the study variables nutritional status, comorbidity, length of hospital stays, presence of polypharmacy satisfied the conditions for multi-variable logistic regression analysis of DRPs. On multivariable logistic regression analysis polypharmacy, prolonged hospital stays, and the presence of comorbidities were the independent predictors of DRPs (Table 7).

**Table 7:**
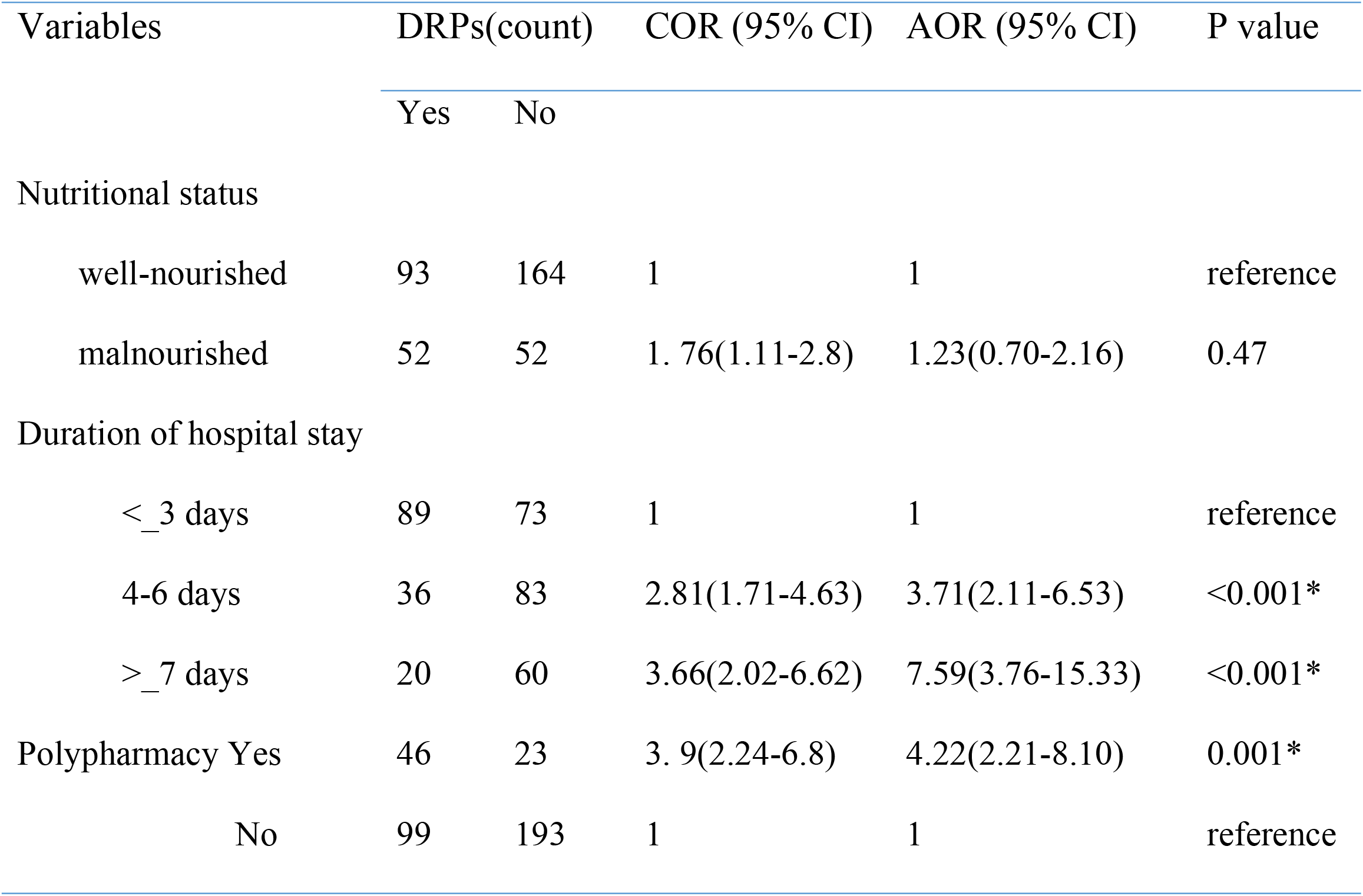
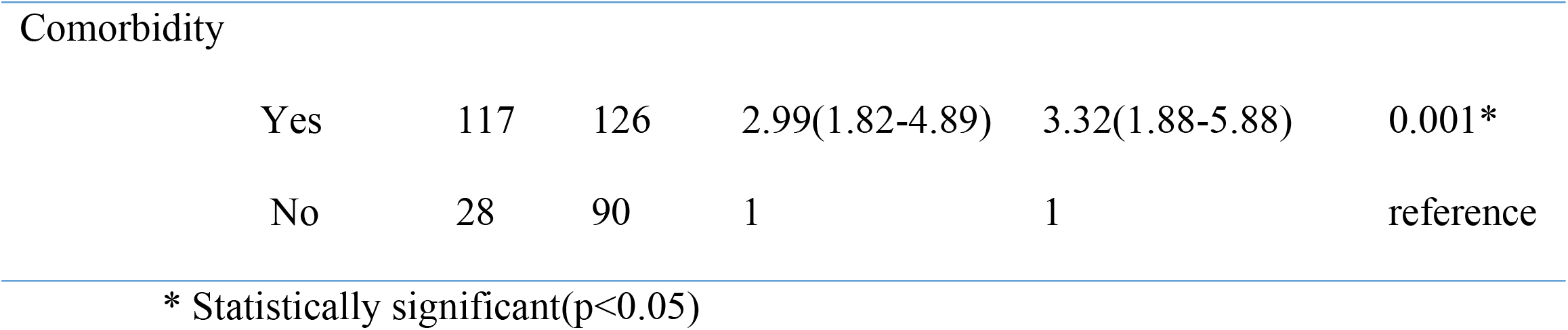
Multivariate logistic regression result of factors associated with DRPs at the pediatric ward of UoGCSH, Ethiopia, May 1 to July 30,2021

| Variables | DRPs(count) |  | COR (95% CI) | AOR (95% CI) | P value |
| --- | --- | --- | --- | --- | --- |
|  | Yes | No |  |  |  |
| Nutritional status |  |  |  |  |  |
| well-nourished | 93 | 164 | 1 | 1 | reference |
| malnourished | 52 | 52 | 1. 76(1.11-2.8) | 1.23(0.70-2.16) | 0.47 |
| Duration of hospital stay |  |  |  |  |  |
| <_3 days | 89 | 73 | 1 | 1 | reference |
| 4-6 days | 36 | 83 | 2.81(1.71-4.63) | 3.71(2.11-6.53) | <0.001* |
| >_7 days | 20 | 60 | 3.66(2.02-6.62) | 7.59(3.76-15.33) | <0.001* |
| Polypharmacy Yes | 46 | 23 | 3. 9(2.24-6.8) | 4.22(2.21-8.10) | 0.001* |
| No | 99 | 193 | 1 | 1 | reference |
| Comorbidity |  |  |  |  |  |
| Yes | 117 | 126 | 2.99(1.82-4.89) | 3.32(1.88-5.88) | 0.001* |
| No | 28 | 90 | 1 | 1 | reference |

\* Statistically significant( $p < 0.05$ )

## Discussion

The finding of this study showed that a significant proportion of hospitalized pediatrics patients experience huge number of DRPs which is a major obstacle to achieve therapeutic outcome.

The current study showed that 40.2% (95% CI 35.5-45.4) pediatric patients had at least one DRP which is comparable to the study in the United Kingdom and Saudi Arabia (45.2%)(41) and India (44%)(42). A higher incidence of DRPs were reported in Dessie (87.7%)(43) and Jimma (74.3%)(20). This difference might be due to the difference in the study design, sample size, and the presence of clinical pharmacists in the pediatric ward. For example, the studies in Jimma and Dessie were cohort while our study was cross-sectional and the sample size in Dessie was 81 while our sample was 361 which might cause lower DRP in this setup. The benefits of pharmacist involvement and active participation also appear significant when directly involved in ward rounds by rapidly identifying medication errors and provide real-time advice and recommendations to prescribers(44, 45). A lower prevalence of DRP was reported in Hong Kong (21%)(46). This difference might be due to the difference in the hospital setting including the composition of healthcare workers, training levels of prescribers, presence of support system. The presence of electronic dispensing and prescribing system reduce DRP prevalence (17, 47, 48).

In this study, the most frequently identified DRPs were dosing problems (56.11%) with dose too low (35.56%) and dose too high (20.55%). Similarly, several studies have also reported dosing problems to be the most frequent DRPs among hospitalized pediatric patients (16, 17, 28, 46). pediatrics suffered from dosing problems more than adults because the drug dose is calculated according to a child’s weight and fractional dosing(49). High prevalence of dosing problems in this study was attributed to wrong drug dose calculation by using patient weight and increasing the number of prescribed drugs which increases drug-drug interactions.

Dosing problems might result in ineffective treatment due to low concentration or toxicity due to overdose which increased hospitalization, cost of treatment, morbidity, and mortality(3). The high prevalence of dosing problems in this study would make this an important area requiring further investigation. The implementation of a standard dosing guide will reduce pediatric dosing errors(50).

The need for the additional drug (28.89%) was the second most common identified DRP which is comparable to a study in Dessie (43) (25.2%). A lower prevalence was reported in Addis Ababa (3%)(17). A higher prevalence was reported in Jimma (34.1%)(18).

This discrepancy might be due to the differences in the study duration, study population and study design. For example, the study in Jimma was conducted for longer period (4 months) included only neonates with sepsis who necessitates different drugs. A study in Addis Ababa was retrospective which doesn’t exactly assess actual patient needs which causes lower prevalence.

In this study, the prevalence of ADR was found to be 6.67% which is comparable with a study in Dessie (8.4%)(43) and Addis Ababa (8.49%). A higher incidence was reported in Brazil (22%)(51). Lower incidence was reported in Jimma (2.81%)(20). These differences could be attributed to genetic, disease pattern, difference in study duration and design (51-53). For example, studies conducted for longer period could detect high adverse drug reactions than others like the study conducted in Brazil for 6 months. In addition, a study in Brazil includes all pediatric patients but cancer patients were excluded in this study which may lower ADR prevalence. A study in Jimma only includes pediatrics diagnosed with infectious disease which might lower overall ADR prevalence.

In the present study, 4.44% of total DRPs were identified as unnecessary drug use which is comparable with a study in Addis Ababa (7.5%)(17). In this era of inflation, drug therapy costs are on the rise. This is a burden for developing nations particularly Ethiopia. Therefore, the prevention of unnecessary drug therapy will contribute to cost savings and other drug-related sufferings among hospitalized pediatric patients.

Anti-infectives (61.7%) were the drug classes most implicated in DRPs and this is supported by studies from Addis Ababa(17) and Saudi Arabia(16). Most patients were diagnosed with infectious diseases and anti-infectives were also prescribed as prophylaxis particularly for patients with SAM(54). This higher utilization of anti-infectives than any other class of drugs may be one reason anti-infectives appears as the major class of drugs responsible for causing DRPs(55).

Clinical pharmacists intervened to resolve the identified DRPs. The dose adjustment was the major intervention performed (52%) which is comparable with a study in Cote d’Ivoire (32%)(19). The addition of drugs was the second most performed intervention (30%) which is comparable with a report in Addis Ababa (31.4%)(28). In contrast to this, the commonest interventions done in Jimma were change the medication and adherence and counselling(20). The discrepancy might be due to the dosing problem was the most common type of DRP in this study, whereas noncompliance was the most common DRP in the study at Jimma.

In this study, 76.5% interventions were accepted. A higher acceptance rate was reported in France (98%)(56) and India (86.6%)(42). These differences might be due to differences in the awareness of the relevance of pharmaceutical interventions performed, an integration of the pharmacist in the health-care team and competency of clinical pharmacists to provide evidence based medicine(19). In this study, the presence of comorbidity was one of the independent predictors of DRP. This is supported by studies in South West Ethiopia(20) and Addis Ababa(17). The odds of developing DRPs were 3.3 times higher among patients having comorbidity compared to those patients without comorbidity. This might be due to the presence of comorbid conditions requiring additional drugs which increases the likelihood of adverse drug effects, drug-drug interactions, and non-adherence which increased the likelihood of experiencing DRPs(57).

The present finding showed that polypharmacy was found to be another independent predictor of DRP which is supported with studies in Hong Kong (46) and London(58). The odds of developing DRPs were 4.2 times higher among patients who took greater than four drugs compared to those patients who took less than five drugs This might be due to polypharmacy increases adverse drug reactions, PDDIs, hospital readmissions, and medical costs and nonadherence(59).

In the present study, participants with prolonged hospital stay were more likely to have DRPs and this is supported by studies done in Vietnam (60) and South West Ethiopia(20). The odds of developing DRPs were 7.5 times higher among patients hospitalized greater than six days compared to those patients who were hospitalized for less than three days. Similarly, the odds of DRPs were 3.7 times higher among patients hospitalized 4 to 6 days compared to those patients who were hospitalized for less than three days. This might be due to prolonged hospitalization may increase poly physician visit having different knowledge about pediatric treatment that increases the no of drugs, drug-drug interactions, increased cost, and non-adherence which increases the risk of DRPs. Another possible reason could be the more the patient stayed in the hospital, the more likely the patient had a chance to acquire new infections such as hospital-acquired infections. These infectious diseases need new medications which further contributed to the occurrence of DRP(43).

## Limitation of the study

First, as we excluded intensive care unit and oncology patients, it may not be possible to generalize the results obtained to the entire pediatric population. The study did not assess the outcome of intervention provided and DRP. The study was conducted at a single institution using cross-sectional study design. Lastly, the study also doesn’t consider the use of medicine in off-label patterns outside the indication in pediatrics as the cause of drug-related problems

## Conclusion

The magnitude of DRP in hospitalized patients at the pediatric ward of UoGCSH was found to be high. Dosing problems and needs additional drugs were the most common identified DRPs among hospitalized pediatric patients at the UoGCSH.

The finding of our study revealed that the presence of comorbidity, polypharmacy and prolonged hospital stays were independent predictors of DRPs.

## Recommendation

Pediatric drug dosing should be done strictly to prevent inappropriate doses. Strategies have to be developed to monitor the performance and the problems encountered by clinical pharmacists. Further follow up and multicenter studies should be done at the pediatric ward to get comprehensive results. Researchers have to research the clinical, economic impact of DRP among pediatric patients admitted to the pediatric ward. Lastly, there should be active participation of clinical pharmacists during the provision of pharmaceutical care with other health care professionals.

## Data Availability

All relevant data are within the manuscript and its Supporting Information files.

## Acknowledgements

The authors would like to extend their gratitude to all individuals involved in data collection and patients who participated in the study.

## Ethical approval

Ethical clearance was obtained from the ethical review committee of department of clinical Pharmacy, Gondar University, and permission was also obtained from University of Gondar Comprehensive and Specialized Hospital and unit officials to conduct this study. Written consent was obtained from caregiver and verbal assent was given to study participants. study participants or caregivers were informed about the purpose of the study and their participation was voluntary.

## Informed consent

Written informed consent and assent was obtained from legally authorized representatives (parents/caregiver) before the study.

## Availability of data and materials

The data sets used and/or analyzed during the current study are available from the corresponding author on reasonable request.

